# Associations of Insomnia Symptoms and Trajectories with Incident Cardiovascular Disease: A Population-Based Cohort Study

**DOI:** 10.1101/2024.09.09.24313365

**Authors:** Qing-Mei Huang, Hao-yu Yan, Huan Chen, Jia-Hao Xie, Jian Gao, Zhi-Hao Li, Chen Mao

**Author notes:** Correspondence to Chen Mao, PhD; Department of Epidemiology, School of Public Health, Southern Medical University, Guangzhou 510515, Guangdong, China. **Qing-Mei Huang and Hao-Yu Yan** contributed equally to this article.

## Abstract

**Background:** There is limited understanding regarding associations between insomnia symptoms, particularly the trajectories of insomnia symptoms, and cardiovascular disease (CVD). We aimed to investigate the associations of insomnia symptoms and trajectories with the risk of incident CVD.

**Methods:** This study used data from the Health and Retirement Study. Insomnia symptoms included non-restorative sleep, difficulty initiating sleep, early morning awakening, and difficulty maintaining sleep, classified on a scale ranging from 0 to 8. We also identified four distinct trajectories of insomnia symptoms: low, decreasing, increasing, and high insomnia symptoms. Examined outcomes included incident heart disease, stroke, and the combination of the two referred as CVD in the present study. Cox proportional hazard models were used to calculate the hazard ratio (HR) and 95% confidence interval (95% CI) after adjusting for potential confounders.

**Results:** A total of 12 102 participants aged 50 years or over without CVD at baseline were included. During a median follow-up of 10.2 years, 3 962 first CVD events occurred (3 372 heart disease and 1 200 stroke). Participants experiencing one (HR, 1.16 [95% CI, 1.05-1.27]), two (HR, 1.16 [ 95% CI, 1.05-1.28]), or three to four (HR, 1.26 [95% CI, 1.15-1.38]) insomnia symptoms had a higher risk of incident CVD compared to those not experiencing any insomnia symptoms. After a median follow-up of 8.4 years after the visit 2, 2 375 first CVD events occurred (1 981 heart disease and 705 stroke). Using the trajectory with low insomnia symptoms as the reference, increasing insomnia symptoms (HR, 1.28 [95% CI, 1.10-1.50]) and high insomnia symptoms (HR, 1.32 [95% CI, 1.15-1.50]) were associated with an increased risk of incident CVD.

**Conclusions:** Higher insomnia symptoms and increasing insomnia symptoms over time are associated with a higher risk of CVD in the community. Public health awareness and screening for insomnia symptoms in the middle-aged and elderly population should be encouraged to reduce CVD.

**Clinical Perspective:** *What Is New:*

- Participants experiencing one, two, or three to four insomnia symptoms had a higher risk of incident CVD compared to those not experiencing any insomnia symptoms.
- Higher insomnia symptoms and increasing insomnia symptoms over time are associated with a higher risk of CVD in the community.

*What Are the Clinical Implications:*

- Early prevention and mitigation of insomnia symptoms could be beneficial in reducing the population burden of CVD.
- Clinicians can provide specific recommendations to improve sleep hygiene as part of a comprehensive approach to reducing the risk of incident CVD.

## Introduction

Cardiovascular disease (CVD) is an ongoing epidemic and a serious clinical and public health issue. In the United States, an estimated 127.9 million adults are living with CVD^1^. CVD is a leading cause of hospital admissions and mortality, with an annual economic burden reaching approximately $407.3 billion^1^. Despite advances in CVD treatment and an enhanced understanding of its risk factors, prognosis and patients’ quality of life remain sub-optimal^1^. Notably, approximately 70% of CVD mortality are attributed to modifiable risk factors^2^. Healthy lifestyle habits, such as maintaining a normal body weight^3^, quitting or not smoking^4^, and engaging in regular exercise^5^ reduce the lifetime risk of CVD. Additionally, sustaining these healthy habits exerts a positive influence, preventing the occurrence of other major risk factors for CVD^4^, including hypertension^6^, obesity^7^, and diabetes^8^. Thus, the identification and targeting of these modifiable factors hold the potential to reduce CVD incidence and improve the overall quality of life.

Insomnia symptoms, such as difficulty falling asleep, difficulty maintaining sleep, waking up in the morning, and non-restorative sleep, are common among middle-aged and older adults^9^. As many as 50% of middle-aged people and up to 75% of older adults report experiencing at least one type of insomnia symptom annually^10^. Mounting evidence has demonstrated the associations between insomnia symptoms and adverse cardiovascular outcomes, including myocardial infarction, stroke, coronary heart disease, and cardiovascular-related deaths^11^. Notably, in 2016, the American Heart Association urged health organizations to formulate evidence-based sleep recommendations for various sleep disorders, encompassing insomnia^12^. However, the definitions of insomnia in most previous cohort studies are not clear enough or solely rely on the symptom of having difficulties initiating sleep, neglecting a comprehensive examination of other insomnia symptoms. Given that different insomnia symptoms correlate with distinct underlying mechanisms and pathological alterations, it is possible that their effects on CVD risks may exhibit significant variations. Limited research has explored the associations between concurrent insomnia symptoms and the risk of incident CVD.

Additionally, numerous previous cohort studies included only a single measurement of insomnia at baseline, ignoring the dynamic feature of insomnia over time. The trajectory of insomnia symptoms exhibits considerable variability among individuals over time. However, the effects of insomnia symptoms trajectories on CVD outcomes remain inadequately investigated, resulting in a loss of life-course exposure information concerning the role of insomnia in CVD etiology. Due to the limited comprehension of prospective associations between insomnia symptoms, especially their trajectories, and CVD, it is imperative to conduct a study that covers a large community-dwelling population with a relatively extended follow-up period to better understand the association between insomnia symptoms, their trajectories, and CVD.

Accordingly, we used longitudinal population-based cohort data from the Health and Retirement Study (HRS) to investigate the associations of insomnia symptoms and the trajectories of insomnia symptoms with CVD.

## Methods

### Study design and participants

The study population and design of the HRS have been previously described^13^. Briefly, the HRS is a population-based cohort of community-dwelling adults in the United States and their spouses of any age. The HRS implemented biennial interviews to acquire detailed health and population data from the point of queue entry until either departure or death. The study obtained approval from the Institutional Review Committee of the University of Michigan and the National Institute on Aging (HUM00061128). Verbal informed consent is obtained from all participants in the HRS before their involvement in the study.

In the current study, participants were from one wave of the HRS (2002) and were limited to those aged 50 or older at baseline (n=17 758). In the insomnia symptoms association analysis, participants with missing data on insomnia symptoms at baseline (2002), those reporting CVD at baseline, or those lost to follow-up at any time point were excluded, leaving 12 102 participants who were included. In the trajectories of insomnia symptoms association analysis, participants with missing data on insomnia symptoms at the three examination rounds in 2002 (baseline), 2004 (visit 1), and 2006 (visit 2), those reporting CVD at the three examination rounds, or those lost to follow-up at any time point were excluded, leaving 8 731 participants who were included (Figure S1 in the Supplement).

### Assessment of insomnia symptoms

Participants were asked about insomnia symptoms at each wave. Questions about insomnia symptoms assessed difficulties in initiating and maintaining sleep, early morning awakening, and non-restorative sleep. Participants were asked how often they have trouble with ‘falling asleep’, ‘waking up during the night’, and ‘waking up too early and not being able to fall asleep again’, and how often they feel ‘really rested’ when they wake up in the morning. The response options for each question included ‘most of the time’, ‘sometimes’, and ‘rarely or never’. We defined participants as experiencing insomnia symptoms when they answered ‘most of the time’ in the first three questions and answered ‘rarely or never’ in the last question, as described by prior studies^14^. Each insomnia symptom status was then represented with a binary variable, ‘yes vs. no’, in the analyses. To examine the quantity of insomnia symptoms, we categorized participants based on the number of reported symptoms. This was performed by summing across the four symptoms and categorizing the respondents into four groups: 1) experiencing no symptoms, 2) experiencing one symptom, 3) experiencing two symptoms, and 4) experiencing three to four symptoms. To examine the trajectories of insomnia symptoms from 2002 to 2006, responses ranged from 2 (‘most of the time’) to 0 (‘rarely or never’) for the first three questions and ranged from 0 (‘most of the time’) to 2 (‘rarely or never’) for the last question. We summed values from the four items to compute an insomnia score for each wave, ranging from 0 to 8^15^.

### Ascertainment of outcomes

The main outcomes of interest included the occurrence of heart disease and stroke, and the combination of the two referred as CVD in the present study. The heart disease and stroke status were ascertained through respondents’ self-report of a physician’s diagnosis at each follow-up wave or from proxy informants’ reports of a physician’s diagnosis during post-mortem exit interviews. Specifically, two separate questions asked participants/proxies to indicate whether a physician ever told them that participants had heart disease (heart attack, coronary heart disease, angina, congestive heart failure, or other heart problems) or stroke at each follow-up wave. The variable indicated 1 if the participants had ever had the condition and 0 if otherwise.

### Covariates

Three variable sets were considered potential confounding factors and were defined as follows. The first set defined demographic variables: age, sex (male or female), ethnicity (non-Hispanic white, non-Hispanic black, Hispanic, or other), education level (less than college or college and above), and body mass index (BMI). The second set included lifestyle factors: current smoking (yes or no), alcohol drinking (yes or no), and regular exercise (yes or no). The third set included self-reported physician-diagnosed diseases: hypertension (yes or no), diabetes (yes or no), cancer (yes or no), and chronic lung disease (yes or no). BMI was equal to weight divided by squared body height in kilograms /m^2^.

### Statistical analysis

The baseline characteristics of the included participants were summarized as numbers (percentages) for categorical variables and as means (SDs) for normally distributed continuous variables. Differences in characteristics between the number of insomnia symptoms or the trajectories of insomnia symptoms were tested using analysis of variance or *χ* ^2^ tests. The multiple imputation method was used to correct the missing values and reduce the possibility of inferential bias.

We first used latent class trajectory models to identify trajectories of insomnia symptoms over time. This is a specialized form of finite mixture modeling and is designed to identify latent classes of participants following similar progressions of a determinant over time^16^. Our models used second-order polynomials. For every participant, we calculated the posterior probabilities for each trajectory, and we assigned participants post hoc to the trajectory with the highest probability. We estimated the best-fitting number of trajectories based on a minimum Bayesian Information Criterion^17^, while maintaining the posterior probabilities by class (>0.70) and class size (≥2% of the population). To facilitate interpretability, we assigned labels to the trajectories based on their modeled graphic patterns.

To assess the risk of incident CVD (including heart disease and stroke), the survival model time zero was the examination date of the first examination round (baseline) in the insomnia symptoms association analysis and the examination date of the third examination round in the trajectories of insomnia symptoms association analysis. Cox proportional hazards regression models were used to assess the associations of insomnia symptoms (the number and type of insomnia symptoms) and the trajectories of insomnia symptoms with the risk of incident CVD (including heart disease and stroke). The hazard ratios (HRs) along with the 95% confidence interval (CI) were calculated. Proportional hazards assumptions were not violated when assessed using Schoenfeld residuals (*P*>0.05). For all analyses, we fitted a multivariate model which was adjusted for age, sex, ethnicity, education level, BMI, current smoking, alcohol drinking, regular exercise, hypertension, diabetes, cancer, and chronic lung disease. Furthermore, sensitivity analyses were performed to exclude participants missing covariate data and to exclude those who developed heart disease or stroke during the first two years of follow-up to account for the possibility of reverse causation.

All statistical analyses were performed in R software, version 4.3.2 (R Project for Statistical Computing). Reported *P* values were two-tailed, and *P* < 0.05 was considered statistically significant.

## Results

### Baseline Characteristics

The baseline characteristics of the study participants according to the number of insomnia symptoms are reported in Table 1. Of the 12 102 participants with a median age of 67.2 years, 61.1% were female. The proportion of those with at least one insomnia symptom was 77.6%. Compared with participants with no insomnia symptom, those with at least one insomnia symptom were more likely to be older, to be female, to be non-Hispanic White, to be less educated, to have a higher BMI, and to have a higher prevalence of hypertension, diabetes, and chronic lung disease. They were less likely to be current smokers or physically active. The characteristics of the study participants according to the trajectories of insomnia symptoms are shown in Table S1 in the Supplement.

**Table 1.**
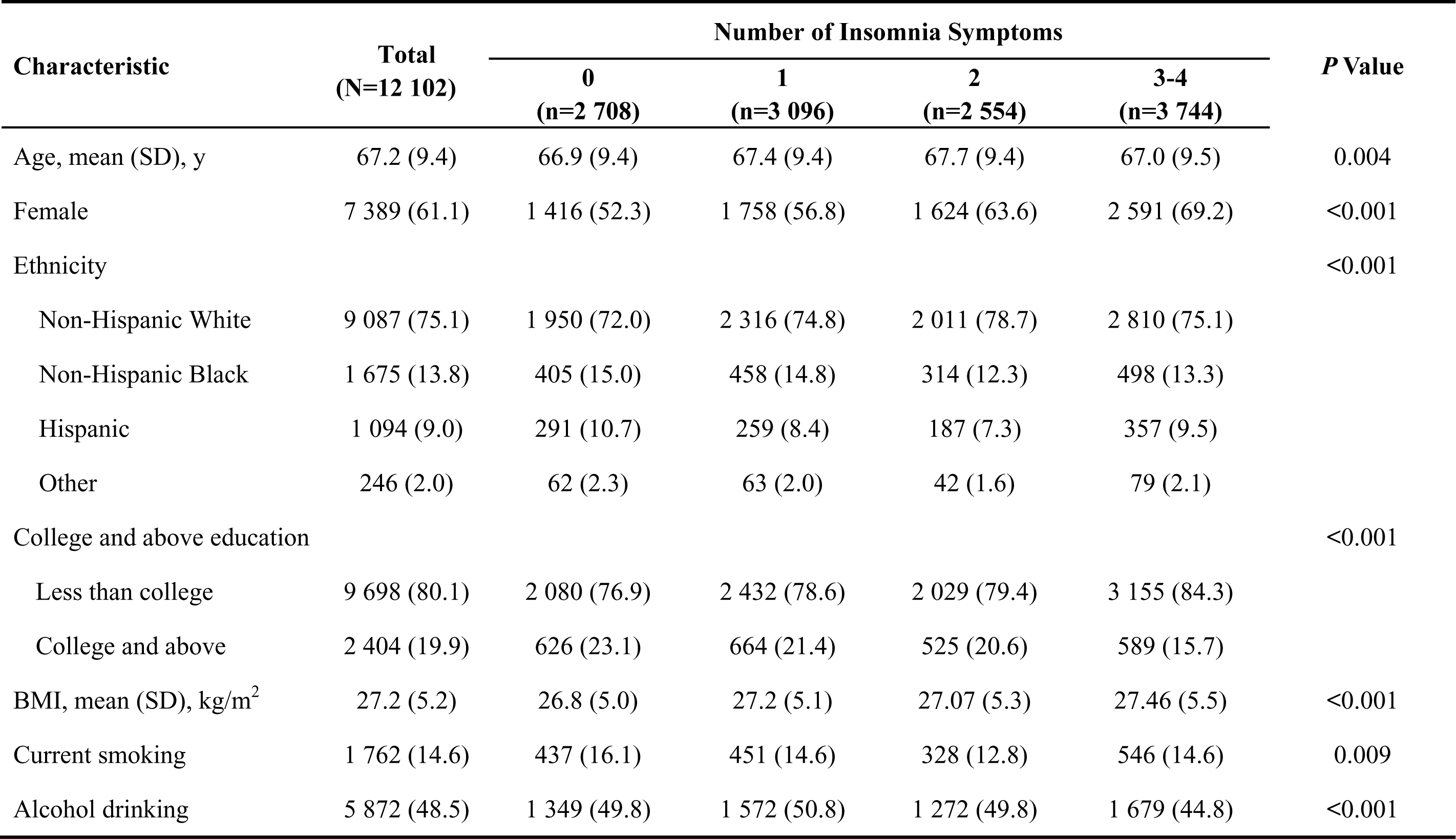
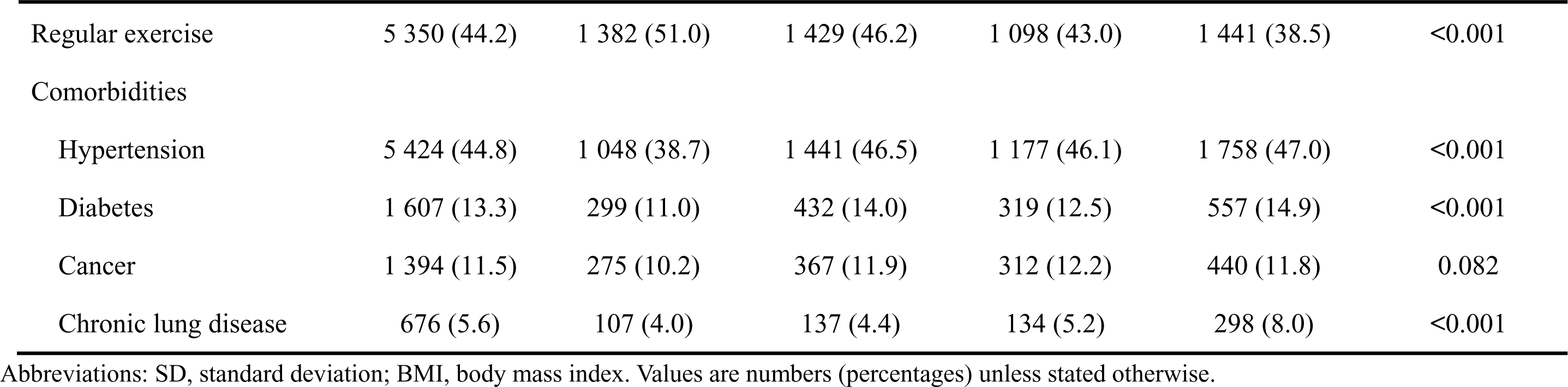
Baseline Characteristics of Study Participants According to Number of Insomnia Symptoms.

| Characteristic | Total<br>(N=12 102) | Number of Insomnia Symptoms |  |  |  | P Value |
| --- | --- | --- | --- | --- | --- | --- |
|  |  | 0<br>(n=2 708) | 1<br>(n=3 096) | 2<br>(n=2 554) | 3-4<br>(n=3 744) |  |
| Age, mean (SD), y | 67.2 (9.4) | 66.9 (9.4) | 67.4 (9.4) | 67.7 (9.4) | 67.0 (9.5) | 0.004 |
| Female | 7 389 (61.1) | 1 416 (52.3) | 1 758 (56.8) | 1 624 (63.6) | 2 591 (69.2) | <0.001 |
| Ethnicity |  |  |  |  |  | <0.001 |
| Non-Hispanic White | 9 087 (75.1) | 1 950 (72.0) | 2 316 (74.8) | 2 011 (78.7) | 2 810 (75.1) |  |
| Non-Hispanic Black | 1 675 (13.8) | 405 (15.0) | 458 (14.8) | 314 (12.3) | 498 (13.3) |  |
| Hispanic | 1 094 (9.0) | 291 (10.7) | 259 (8.4) | 187 (7.3) | 357 (9.5) |  |
| Other | 246 (2.0) | 62 (2.3) | 63 (2.0) | 42 (1.6) | 79 (2.1) |  |
| College and above education |  |  |  |  |  | <0.001 |
| Less than college | 9 698 (80.1) | 2 080 (76.9) | 2 432 (78.6) | 2 029 (79.4) | 3 155 (84.3) |  |
| College and above | 2 404 (19.9) | 626 (23.1) | 664 (21.4) | 525 (20.6) | 589 (15.7) |  |
| BMI, mean (SD), kg/m<sup>2</sup> | 27.2 (5.2) | 26.8 (5.0) | 27.2 (5.1) | 27.07 (5.3) | 27.46 (5.5) | <0.001 |
| Current smoking | 1 762 (14.6) | 437 (16.1) | 451 (14.6) | 328 (12.8) | 546 (14.6) | 0.009 |
| Alcohol drinking | 5 872 (48.5) | 1 349 (49.8) | 1 572 (50.8) | 1 272 (49.8) | 1 679 (44.8) | <0.001 |
| Regular exercise | 5 350 (44.2) | 1 382 (51.0) | 1 429 (46.2) | 1 098 (43.0) | 1 441 (38.5) | <0.001 |
| Comorbidities |  |  |  |  |  |  |
| Hypertension | 5 424 (44.8) | 1 048 (38.7) | 1 441 (46.5) | 1 177 (46.1) | 1 758 (47.0) | <0.001 |
| Diabetes | 1 607 (13.3) | 299 (11.0) | 432 (14.0) | 319 (12.5) | 557 (14.9) | <0.001 |
| Cancer | 1 394 (11.5) | 275 (10.2) | 367 (11.9) | 312 (12.2) | 440 (11.8) | 0.082 |
| Chronic lung disease | 676 (5.6) | 107 (4.0) | 137 (4.4) | 134 (5.2) | 298 (8.0) | <0.001 |

Abbreviations: SD, standard deviation; BMI, body mass index. Values are numbers (percentages) unless stated otherwise.

### Insomnia Symptoms and the Risk of Incident CVD

During a median follow-up of 10.2 years, a total of 3 962 first CVD events occurred including 3 372 heart disease and 1 200 stroke. The associations of the cumulative number and type of insomnia symptoms with the risk of incident CVD, heart disease and stroke are reported in Table 2. After adjusting for multiple confounding variables, participants experiencing one (HR, 1.16 [95% CI, 1.05-1.27]), two (HR, 1.16 [95% CI, 1.05-1.28]), or three to four (HR, 1.26 [95% CI, 1.15-1.38]) insomnia symptoms had a higher risk of incident CVD compared to those not experiencing any insomnia symptoms. For each symptom individually, experiencing non-restorative sleep (HR, 1.11 [95% CI: 1.04-1.19]), difficulty initiating sleep (HR, 1.12 [95% CI, 1.05-1.20]), or difficulty maintaining sleep (HR, 1.15 [95% CI, 1.08-1.23]) was associated with a higher risk of incident CVD compared to those not experiencing the symptom.

**Table 2.** Association Between the Cumulative Number and Type of Insomnia Symptoms and CVD Among 12 102 Participants.

| Respondent characteristics | Total | CVD |  | Heart Disease |  | Stroke |  |
| --- | --- | --- | --- | --- | --- | --- | --- |
|  |  | (Heart Disease or Stroke) |  |  |  |  |  |
|  |  | Events | HR (95% CI) | Events | HR (95% CI) | Events | HR (95% CI) |
| Number of Insomnia Symptoms (ref: no symptoms) |  |  |  |  |  |  |  |
| 1 | 3 096 | 1 032 | 1.16 (1.05-1.27) | 873 | 1.16 (1.05-1.28) | 311 | 1.08 (0.91-1.28) |
| 2 | 2 554 | 844 | 1.16 (1.05-1.28) | 731 | 1.20 (1.08-1.34) | 231 | 1.00 (0.84-1.20) |
| 3-4 | 3 744 | 1 321 | 1.26 (1.15-1.38) | 1 125 | 1.26 (1.15-1.40) | 424 | 1.23 (1.05-1.45) |
| Individual Insomnia Symptoms |  |  |  |  |  |  |  |
| Non-restorative sleep (ref: no) | 4 323 | 1 464 | 1.11 (1.04-1.19) | 1 266 | 1.13 (1.05-1.22) | 443 | 1.09 (0.97-1.23) |
| Difficulty initiating sleep (ref: no) | 4 651 | 1 611 | 1.12 (1.05-1.20) | 1 362 | 1.11 (1.03-1.19) | 523 | 1.21 (1.08-1.36) |
| Early morning awakening (ref: no) | 4 800 | 1 627 | 1.04 (0.98-1.11) | 1 382 | 1.04 (0.97-1.11) | 506 | 1.06 (0.95-1.19) |
| Difficulty maintaining sleep (ref: no) | 7 219 | 2 516 | 1.15 (1.08-1.23) | 2 168 | 1.19 (1.11-1.28) | 731 | 1.00 (0.89-1.12) |

Abbreviations: HR, hazard ratio; CI, confidence interval; CVD, cardiovascular disease.
All models adjusted for age, sex, ethnicity, education level, BMI, current smoking, alcohol drinking, regular exercise, hypertension, diabetes, cancer, and chronic lung disease.

### Trajectories of Insomnia Symptoms and the Risk of Incident CVD

This analysis was conducted in 8 731 participants free of CVD at the three examination rounds. As shown in Figure 1, we identified four distinct trajectories of insomnia symptoms as follows (n, [%]): maintained a low insomnia symptom score throughout the follow-up (‘low insomnia symptoms’; 4915 [56.3%]); had a moderately high starting score but then remitted (‘decreasing insomnia symptoms’; 2362 [27.1%]); had a low starting score that steadily increased throughout the follow-up (‘increasing insomnia symptoms’; 626 [7.2%]); and maintained a high score throughout (‘high insomnia symptoms’; 828 [9.5%]). During a median follow-up of 8.4 years, a total of 2 375 first CVD events occurred including 1 981 heart disease and 705 stroke. The associations of the trajectories of insomnia symptoms with CVD, heart disease and stroke are reported in Table 3. Using the trajectory with low insomnia symptoms as the reference, increasing insomnia symptoms (HR, 1.28 [95% CI, 1.10-1.50]) and high insomnia symptoms (HR, 1.32 [95% CI, 1.15-1.50]) were associated with an increased risk of incident CVD. We observed similar patterns of results for heart disease and stroke, but with slightly stronger effect estimates. For example, the hazard ratio for heart disease was 1.36 (1.15 to 1.60) for increasing trajectory and 1.33 (1.15 to 1.54) for high trajectory. The hazard ratio for stroke was 1.35 (1.02 to 1.77) for increasing trajectory and 1.38 (1.08 to 1.75) for high trajectory.

**Figure 1:**
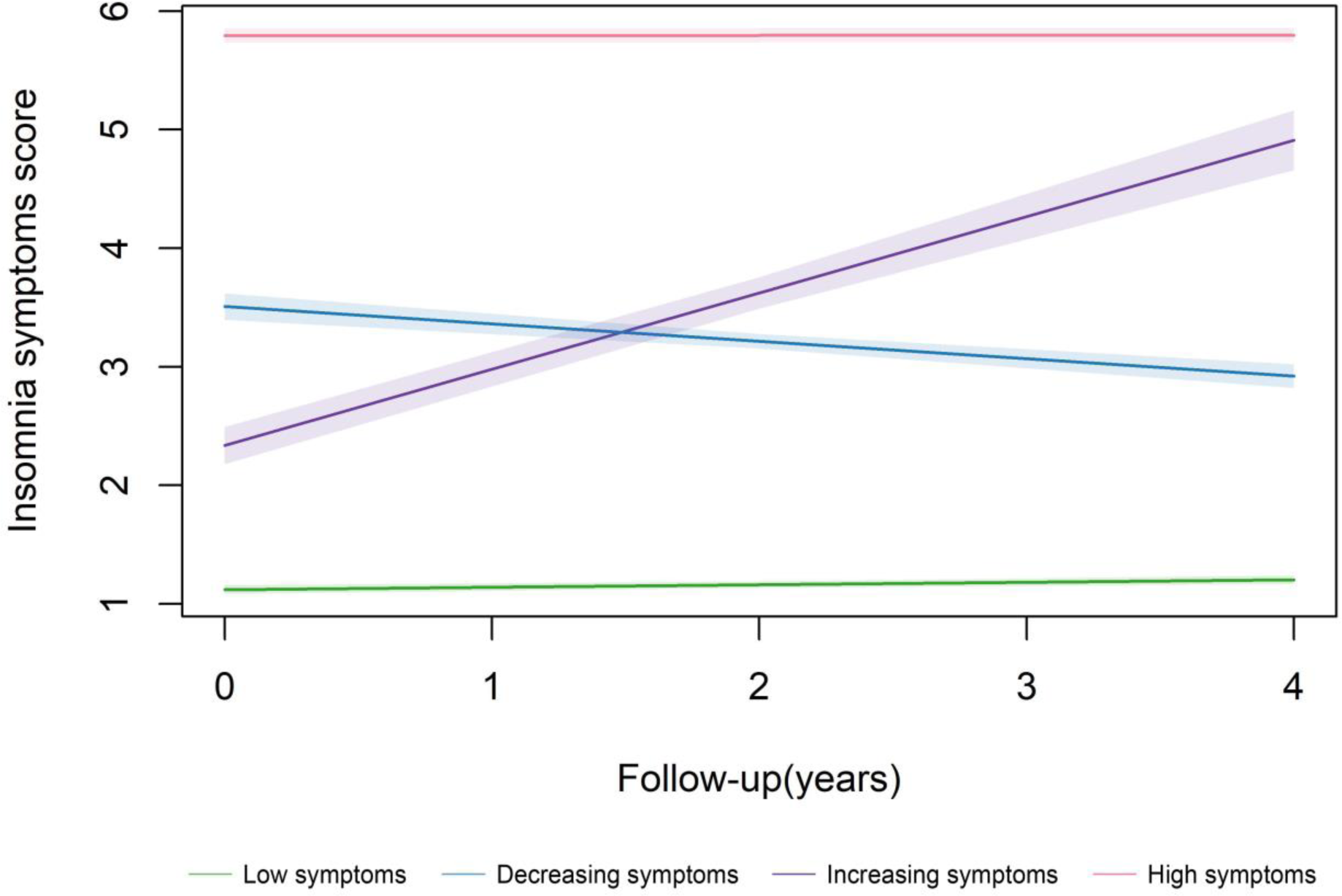
Trajectories of Insomnia Symptoms From 2002–2006. The figure shows trajectories of standardized insomnia symptoms scores over 4 years from 8 731 individuals, with four measures of insomnia symptoms. Shading around the lines represents confidence bands for the calculated trajectory.

**Table 3.** Association Between the Trajectories of Insomnia Symptoms and CVD Among 8 731 Participants.

| Trajectories of Insomnia Symptoms | Total | CVD |  | Heart Disease |  | Stroke |  |
| --- | --- | --- | --- | --- | --- | --- | --- |
|  |  | (Heart Disease or Stroke) |  |  |  |  |  |
|  |  | Events | HR (95% CI) | Events | HR (95% CI) | Events | HR (95% CI) |
| Low | 4 915 | 1 246 | 1.00 (reference) | 1 031 | 1.00 (reference) | 356 | 1.00 (reference) |
| Decreasing | 2 362 | 661 | 1.08 (0.98-1.19) | 553 | 1.09 (0.98-1.21) | 201 | 1.11 (0.93-1.32) |
| Increasing | 626 | 192 | 1.28 (1.10-1.50) | 167 | 1.36 (1.15-1.60) | 61 | 1.35 (1.02-1.77) |
| High | 828 | 273 | 1.32 (1.15-1.50) | 230 | 1.33 (1.15-1.54) | 87 | 1.38 (1.08-1.75) |

Abbreviations: HR, hazard ratio; CI, confidence interval; CVD, cardiovascular disease.
All models adjusted for age, sex, ethnicity, education level, BMI, current smoking, alcohol drinking, regular exercise, hypertension, diabetes, cancer, and chronic lung disease.

### Sensitivity analyses

Sensitivity analysis results are shown in the Table S2-5 in the Supplement. The association of insomnia symptoms and trajectories with CVD, heart disease and stroke was robust and stable in all sensitivity analyses. Similar findings were observed when excluding participants with missing covariates data or excluding participants who developed CVD during the first two years of follow-up.

## Discussion

In this large, nationally representative prospective study of adults in the United States, we found that participants who experienced one or more insomnia symptoms had a higher risk of incident CVD. Furthermore, we identified four distinct trajectories of insomnia symptoms, characterized by low, decreasing, increasing, and high. The trajectories characterized by increasing and high insomnia symptoms were associated with a higher risk of incident CVD.

Our study identified associations of insomnia symptoms with CVD, which were consistent with findings from a number of previous studies. For example, a meta-analysis of 17 cohort studies in middle-aged and older adults reported that insomnia is associated with an increased risk of developing CVD^18^. Another meta-analysis of 13 cohort studies also found that insomnia is associated with an increased risk of CVD^19^. However, the comparability across those studies was compromised by the inconsistency in the definitions of insomnia. A previous study of 487 200 adults aged 30 to 79 years in China found that self-reported difficulties in initiating or maintaining sleep, early morning awakening, and daytime dysfunction were associated with increased risk of CVD (ischemic heart disease and ischemic stroke but not hemorrhagic stroke)^20^ while our findings noted that non-restorative sleep, difficulty initiating sleep and maintaining sleep were associated with a higher risk of incident CVD.

To the best of our knowledge, except for a few studies that conducted repeated time measurements^21^, the main limitation of previous epidemiological studies on sleep and CVD outcomes was single time point measurements. There is evidence to suggest that sleep is a dynamic process that changes during critical periods of aging, but also varies with exposure to various stressors and medications. It is generally accepted that sleep quality worsens in old age^22–24^, but there is heterogeneity in changes in sleep problems over time. Thus, using a latent class trajectory model, four distinct insomnia symptom trajectories were identified in our study based on changes in participants’ insomnia symptoms using 3-time points over 4 years. We noted that participants with increasing and high insomnia symptoms had a significantly higher incidence of CVD. Similarly, a longitudinal study showed that constantly high symptoms had an increased risk of stroke (HR, 1.42 [95% CI: 1.22-1.64]) compared to constantly no symptom^17^.

Some mechanisms underlying this relationship may support our finding, although the pathogenesis of insomnia and CVD is not fully understood. Hypothalamus pituitary axis (HPA) imbalance may be the potential mechanism between insomnia and CVD^25–29^. There is evidence to suggest that the secretion of corticosteroids and cortisol in patients with insomnia increases, especially in patients with short sleep periods, indicating an increase in HPA axis activity. This stress response is also accompanied by increased heart rate, decreased heart rate variability, and increased blood pressure, as well as the secretion of pro-inflammatory cytokines and catecholamines, which are strong risk factors for CVD. What’s more, abnormal regulation of the autonomic nervous system, increased systemic inflammation^30^, increased sympathetic nervous system activity^31^, and increased atherosclerosis^32^ also play an important role in the relationship between insomnia and the risk of CVD. Meanwhile, insomnia is highly comorbid with many mental illnesses. Although insomnia is sometimes considered secondary to depression or other emotional disorders, there is also evidence that insomnia may lead to depression, which can independently contribute to CVD^33^.

Our study has several limitations. Firstly, insomnia symptoms were self-reported, representing subjective indicators of sleep. While technologies such as actigraphy and polysomnography offer precise supplementary sleep data^34^, measures derived from these devices are comparatively less sensitive and specific in recognizing insomnia symptoms. Moreover, their application becomes impractical in extensive epidemiological investigations. Despite the potential for bias in subjective sleep perceptions, they are one of the only means of capturing insomnia symptoms^35^. Secondly, the survey assesses heart disease and stroke as well as other health conditions, primarily relying on self-reported diagnoses from medical professionals. Nevertheless, despite the reliance on self-reporting, these measures have been validated for precise diagnoses of health conditions, particularly CVD, and have consistently shown reliability across diverse populations in previous scientific studies^36^. Finally, this study only utilized data from the HRS dataset. Assessments of sleep duration and other secondary insomnia complaints, such as excessive daytime sleepiness, irritability, and fatigability, were absent in the dataset. Incorporating these variables could have significantly enhanced the depth and comprehensiveness of our analyses.

## Conclusion

In this large population-based study, more insomnia symptoms were associated with an increased risk of CVD. Moreover, the trajectories characterized by high insomnia symptoms and increasing insomnia symptoms were consistently associated with a higher risk of incident CVD. Taken together, these findings suggest that increased awareness and management of insomnia symptoms would likely contribute to preventing CVD occurrence.

## Data Availability

The study obtained approval from the Institutional Review Committee of the University of Michigan and the National Institute on Aging (HUM00061128). Verbal informed consent is obtained from all participants in the HRS before their involvement in the study.

## Non-standard Abbreviations and Acronyms

BMI: BMI, body mass index
CI: confidence interval
CVD: cardiovascular disease
HR: hazard ratio
HRS: Health and Retirement Study
HPA: Hypothalamus pituitary axis
SD: standard deviation

## Acknowledgements

The authors express their gratitude to the participants and staff involved in data collection and management in the Health and Retirement Study.

## Declarations

### Supplemental Material

Tables S1–S5

Figure S1

### Data availability

HRS data are available to researchers by application (https://hrs.isr.umich.edu/).

---

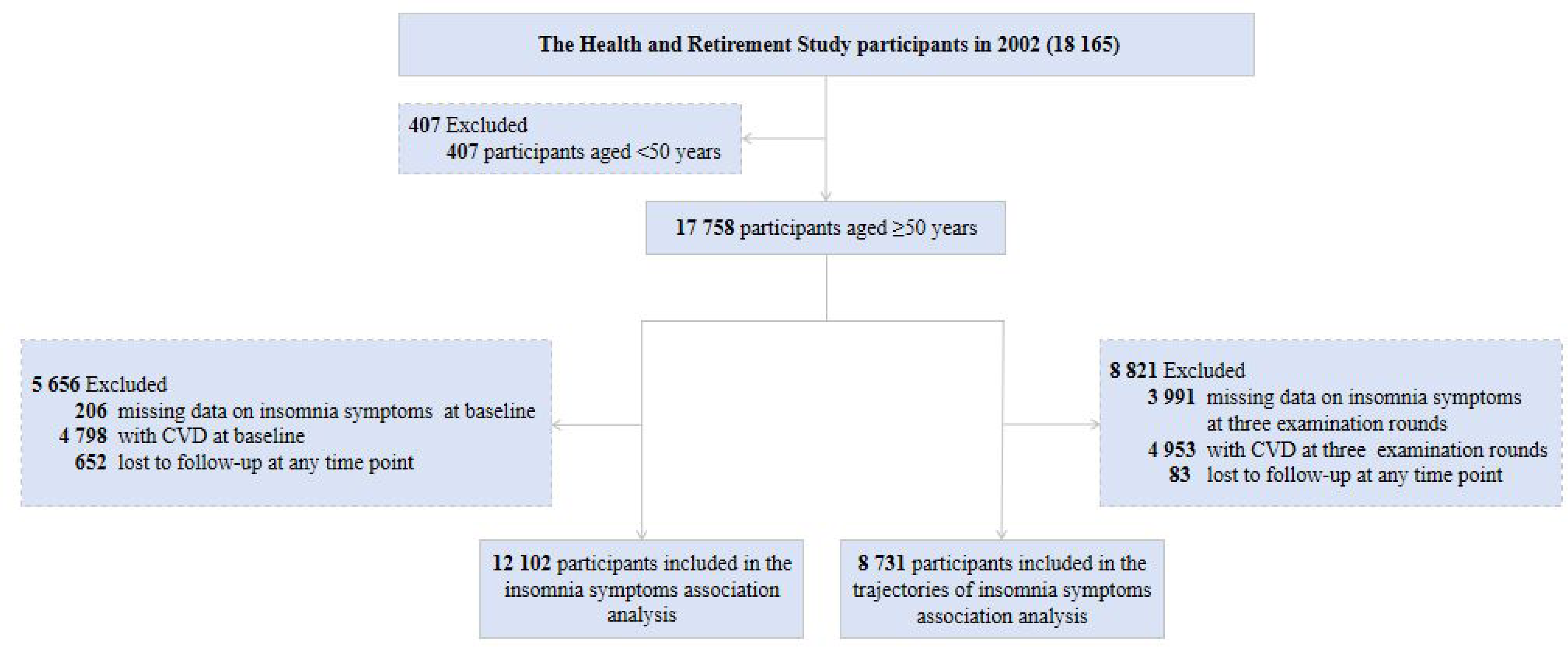

